# Identification of host-pathogen-disease relationships using a scalable Multiplex Serology platform in UK Biobank

**DOI:** 10.1101/19004960

**Authors:** Alexander J Mentzer, Nicole Brenner, Naomi Allen, Thomas J Littlejohns, Amanda Y Chong, Adrian Cortes, Rachael Almond, Michael Hill, Simon Sheard, Gil McVean, UKB Infection Advisory Board, Rory Collins, Adrian VS Hill, Tim Waterboer

## Abstract

**Background:** Certain infectious agents are recognised causes of cancer and potentially other chronic diseases. Identifying associations and understanding pathological mechanisms involving infectious agents and subsequent chronic disease risk will be possible through measuring exposure to multiple infectious agents in large-scale prospective cohorts such as UK Biobank.

**Methods:** Following expert consensus we designed a Multiplex Serology platform capable of simultaneously measuring quantitative antibody responses against 45 antigens from 20 infectious agents implicated in non-communicable diseases, including human herpes, hepatitis, polyoma, papilloma, and retroviruses, as well as *Chlamydia trachomatis, Helicobacter pylori* and *Toxoplasma gondii*. This panel was assayed in a random subset of UK Biobank participants (n=9,695) to test associations between infectious agents and recognised demographic and genetic risk factors and disease outcomes.

**Findings:** Seroprevalence estimates for each infectious agent were consistent with those expected from the literature. The data confirmed epidemiological associations of infectious agent antibody responses with sociodemographic characteristics (e.g. lifetime sexual partners with *C, trachomatis*; *P*=1·8×10^−149^), genetic variants (e.g. rs6927022 with Epstein-Barr virus (EBV) EBNA1 antibodies, *P*=9·5×10^−91^) and disease outcomes including human papillomavirus-16 seropositivity and cervical intraepithelial neoplasia (odds ratio 2·28, 95% confidence interval 1·38-3·63), and quantitative EBV viral capsid antigen responses and multiple sclerosis through genetic correlation (MHC r_G_=0·30, *P=*0·01).

**Interpretation:** This dataset, intended as a pilot study to demonstrate applicability of Multiplex Serology in epidemiological studies, is itself one of the largest studies to date covering diverse infectious agents in a prospective UK cohort including those traditionally under-represented in population cohorts such as human immunodeficiency virus-1 and *C. trachomatis*. Our results emphasise the validity of our Multiplex Serology approach in large-scale epidemiological studies opening up opportunities for improving our understanding of host-pathogen-disease relationships. These data are available to researchers interested in examining the relationship between infectious agents and human health.

## Introduction

Some bacterial, viral and parasitic infectious agents are well established causal factors for certain types of cancer^1^ or have been linked to the development of inflammatory diseases such as multiple sclerosis^2^, and cardiovascular disease^3,4^. However, much of this evidence is derived from cross-sectional or case-control analyses that are unable to determine temporality, or from small nested case-control studies that have generally been small or have focused on a small number of infectious agents, leading to inconsistent findings^5^.

UK Biobank (UKB) is a large prospective cohort study that has collected a large amount of genetic, lifestyle, and biomarker data alongside a wide range of health outcomes for over 500,000 adults in the United Kingdom^6^. The depth and breadth of data available in UKB alongside repeated biological sampling in random subsets every few years offer unique opportunities to explore the inter-relationships between risk factors and disease outcomes in very large numbers of individuals. The availability of serological data characterising lifetime exposure history for multiple infectious agents would allow the retesting of putative and identification of novel associations between infectious agents and multiple disease outcomes whilst accounting for temporal exposure and confounders. Moreover, since many infectious agents such as Epstein-Barr virus (EBV), cytomegalovirus (CMV) and human immunodeficiency virus (HIV-1) are known to have modulatory effects on the immune system, it is equally important to consider their exposure history when testing other non-infectious exposure and disease risk associations. Understanding the role of infectious agents in the development of non-communicable diseases (NCDs) could have major implications for guiding public health decisions such as targeted vaccination strategies for primary prevention^7^.

Here, we describe the results from the first phase of an intended project to measure antibody responses against 45 antigens from 20 infectious agents using a validated Multiplex Serology panel for all 500,000 UKB participants. Using a randomly selected subset of 9,695 individuals, we demonstrate the applicability of the assay platform by confirming expected seroprevalence estimates, and reproducing previously reported epidemiological and genetic associations with infectious agent exposure, highlighting the significant value of these data in understanding the role of infectious agents in the development of NCDs.

## Methods

### Infectious agent selection

A UK Biobank Infectious Disease Working Group was established to provide a consensus list of infectious agents deemed to be of significant importance to public health (**Supplementary Methods**), and to determine the most appropriate methodology for their measurement in a large-scale prospective study (**Supplementary Table 1**). The final Multiplex Serology panel measured antibody responses against 45 antigens spanning 20 infectious agents (**Supplementary Table 2**) including herpes simplex viruses 1 and 2 (HSV-1 and −2); varicella zoster virus (VZV); EBV; CMV; human herpesviruses 6A, 6B and 7 (HHV-6A, HHV-6B and HHV-7); Kaposi’s sarcoma-associated herpesvirus (KSHV); HIV-1; human T-lymphotropic virus-1 (HTLV-1); hepatitis B (HBV) and hepatitis C (HCV) viruses; human papillomaviruses 16 (HPV-16) and 18 (HPV-18); the JC (JCV), BK (BKV) and Merkel cell (MCV) polyomaviruses; *Helicobacter pylori* (*Hp*); *Chlamydia trachomatis* (*Ct*); and *Toxoplasma gondii* (*Tg*).

**Table 1:** Baseline characteristics of the 9,695 randomly selected individuals with Multiplex Serology data compared with the remainder of the UKB cohort.

| Characteristic (range where applicable) | Multiplex Serology<br>measures,<br>Numbers (%)<br>N=9,695 | Remaining UKB<br>Cohort,<br>Numbers (%)*<br>N=493,852 |
| --- | --- | --- |
| **Age at recruitment** |  |  |
| 40-49 | 2543 (26.2) | 129,899 (26.3) |
| 50-59 | 3390 (35.0) | 174,181 (35.3) |
| 60-69 | 3762 (38.8) | 189,765 (38.4) |
| **Sex** |  |  |
| Male | 4271 (44.1) | 225,064 (45.6) |
| Female | 5424 (55.9) | 268,788 (54.4) |
| **Ethnic group** |  |  |
| White | 9140 (94.3) | 464,606 (94.1) |
| Asian | 236 (2.4) | 11238 (2.3) |
| Black | 141 (1.5) | 7877 (1.6) |
| Other | 134 (1.4) | 7390 (1.5) |
| Not reported / missing | 44 (0.4) | 2741 (0.5) |
| **Townsend deprivation index<sup>†</sup>** |  |  |
| Less than -2 (more affluent) | 5106 (52.7) | 259,869 (51.7) |
| -2 to 2 (average) | 3046 (31.4) | 159,450 (31.7) |
| Greater than 2 (more deprived) | 1533 (15.8) | 82343 (16.4) |
| Not reported / missing | 10 (0.1) | 881 (0.2) |
| **Smoking status** |  |  |
| Never | 5377 (55.5) | 268,842 (54.5) |
| Previous | 3308 (34.1) | 170,052 (34.4) |
| Current | 956 (9.8) | 52048 (10.5) |
| Not reported / missing | 54 (0.6) | 2910 (0.6) |
| **Alcohol drinker status** |  |  |
| Never | 425 (4.4) | 21976 (4.4) |
| Previous | 338 (3.5) | 17819 (3.6) |
| Current | 8913 (91.9) | 452,424 (91.7) |
| Not reported / missing | 19 (0.2) | 1633 (0.3) |
| **Number of sexual partners** |  |  |
| 0 | 89 (0.9) | 4148 (0.8) |
| 1 | 2338 (24.1) | 114,636 (23.2) |
| 2-4 | 2534 (26.1) | 128,428 (26.0) |
| 5-10 | 2058 (21.2) | 103,251 (20.9) |
| > 10 | 1007 (10.4) | 51751 (10.5) |
| Not reported / missing | 1669 (17.3) | 91638 (18.6) |
| **Ever had same sex intercourse** |  |  |
| Yes | 295 (3.0) | 15819 (3.1) |
| No | 8442 (87.1) | 437,256 (87.0) |
| Not reported / missing | 958 (9.9) | 49468 (9.9) |

\*: UKB participants with >100 µL serum
†: Townsend deprivation index presented here in categories of integers for representation purposes but modelled in quintiles

**Table 2:**
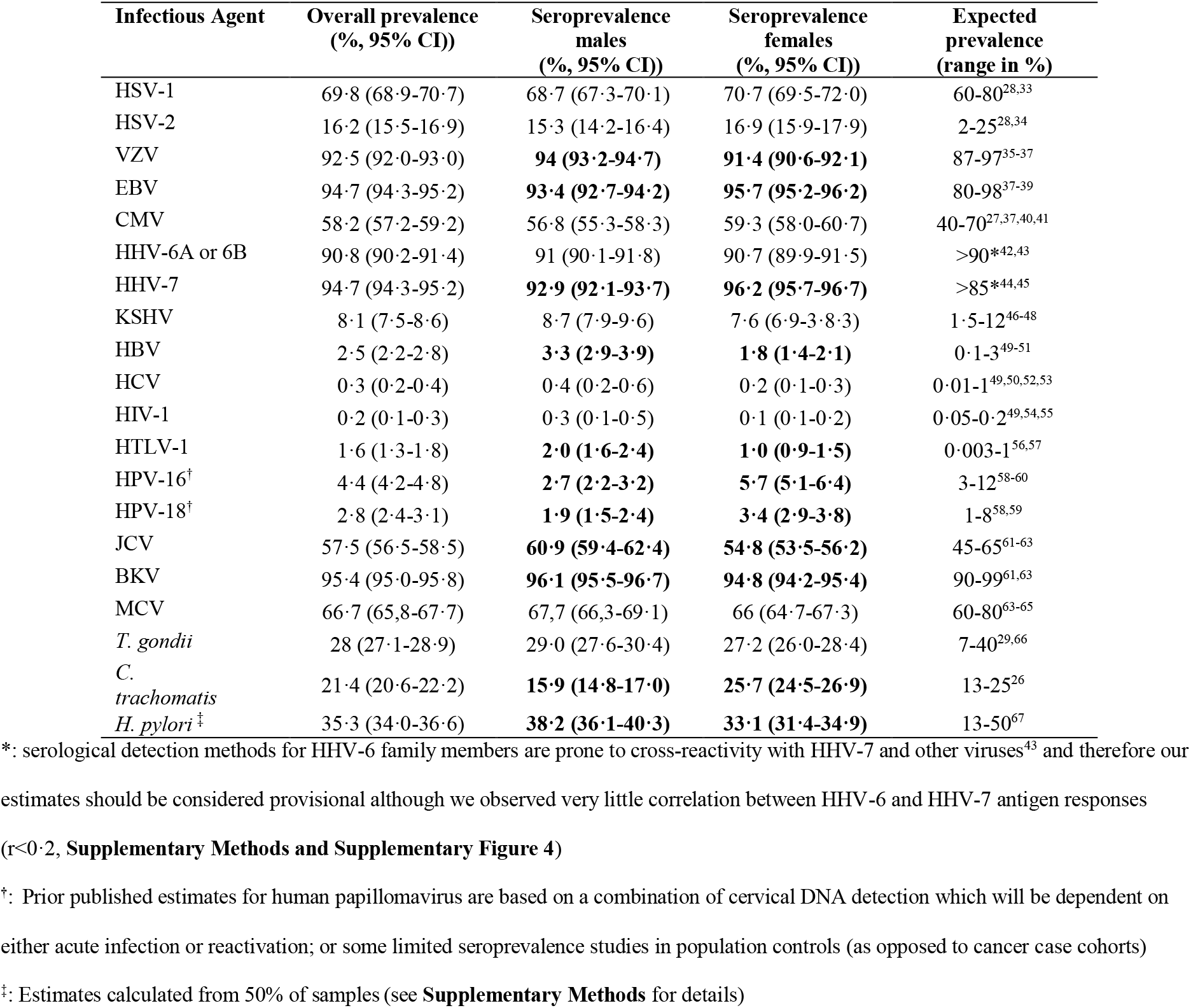
Unadjusted seroprevalence estimates and 95% confidence intervals for infectious agents tested using the Multiplex Serology platform in all 9,695 individuals, overall and stratified by sex. Estimates in men and women highlighted in bold are significantly different from each other (i.e. they do not have overlapping confidence intervals). Ranges of previously published seroprevalence estimates representing expectations are provided as a reference.

### UK Biobank study population

The UKB recruitment process has been described previously^8^. Briefly, half a million men and women aged 40-69 years attended one of 22 UKB assessment centres located throughout England, Scotland and Wales between 2006 and 2010. All participants completed a touchscreen questionnaire, verbal interview and had a range of physical measurements and blood, urine and saliva samples taken for long-term storage. A subset of 20,000 individuals attended a repeat assessment between 2012 and 2013.

For this study, serum samples from 9,695 UKB participants were selected at random and assayed using the final Multiplex Serology panel. For 277 (2·9%) of these participants, an additional sample from the repeat assessment was assayed to assess stability of seroconversion over a 4-5 year period (**Supplementary Methods**).

### Infectious agent assay validation and quality control

Our assay was designed using the principles of an enzyme linked immunosorbent assay (ELISA) that measures levels of serum immunoglobulin G (IgG), which is a stable biological marker of lifetime exposure for a given infectious agent. Unlike ELISAs for individual infectious agents, we implemented a bead-based immunoassay protocol that allows simultaneous measurement of antibodies against multiple agent-specific antigens in the same reaction vessel hence reducing cost and workload. As opposed to many established standard ELISAs, which are predominantly based on inactivated multi-antigenic components of the target infectious agent, our assay is based on measurement of antibody responses against pre-defined recombinantly expressed agent-specific antigens creating opportunities for agent lifecycle-specific antigen characterisation. For each infectious agent tested, we measured antibody responses for up to six antigens. Where two or more antigens were used, we defined algorithms to combine the responses against multiple antigens and designate overall seropositivity or seronegativity against individual infectious agents (**Supplementary Methods**).

The workflow for validation was in line with the criteria outlined in the STARD guidelines^9^ (**Supplementary Figure 1**). The description and validation of the assays for the individual infectious agents described here have been detailed elsewhere (**Supplementary Methods and Supplementary Table 2**)^10-17^. In brief, Multiplex Serology is a bead-based glutathione *S*-transferase (GST) capture assay incorporating glutathione-casein coated fluorescence-labelled polystyrene beads and pathogen-specific GST-X-tag fusion proteins as antigens^17,18^. Full length or fragment viral, bacterial or parasitic antigens were expressed in *E. coli* fused to an N-terminal GST domain and a C-terminal peptide tag for detection of full-length expression and stored as cleared lysate^10-12,14,16,19^. Each antigen was loaded onto one distinct bead set via *in situ* affinity purification of GST fusion proteins on glutathione-derivatised beads. Subsequently, the bead sets (i.e. antigens) were mixed and simultaneously presented to primary serum antibodies (at serum dilution 1:1000). Formed immunocomplexes were detected using a biotinylated goat-α-human IgG secondary antibody and quantified in a Luminex 200 flow cytometer via streptavidin-R-phycoerythrin as reporter dye. Per bead set, at least 100 beads were measured and Median Fluorescence Intensities (MFI) were calculated.

**Figure 1:**
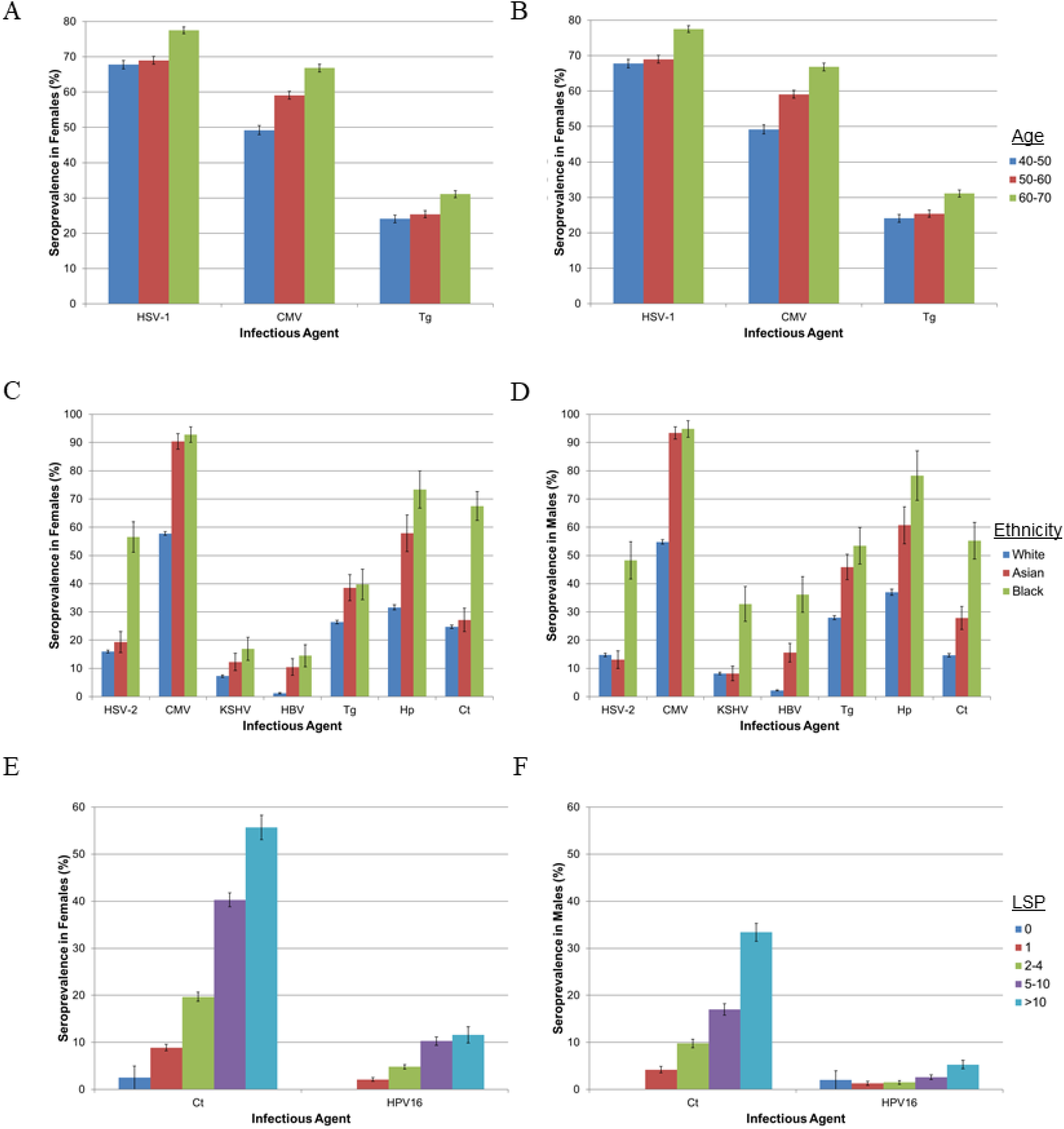
Crude seroprevalence estimates and 95% confidence intervals for multiple infectious agents in 9,695 UK Biobank participants stratified by age (A for females and B for males), self-reported ethnicity (C for females and D for males) and LSP (E for females and F for males).

Individual infectious agent immunoassays were validated by comparing seropositivity and seronegativity estimates from the Multiplex Serology platform against gold standard assays using collections of anonymised, unlinked samples available as reference panels^10-12,14,16^. These panels were also used to ensure that the performance of the agent-specific Multiplex Serology assays conducted on non-magnetic beads were equivalent when combined in the final Multiplex Serology panel tested on magnetic beads (**Supplementary Methods, Supplementary Figure 2 and Supplementary Table 3)**. Magnetic beads are required for full scalability of the Multiplex Serology assay.

**Table 3:**
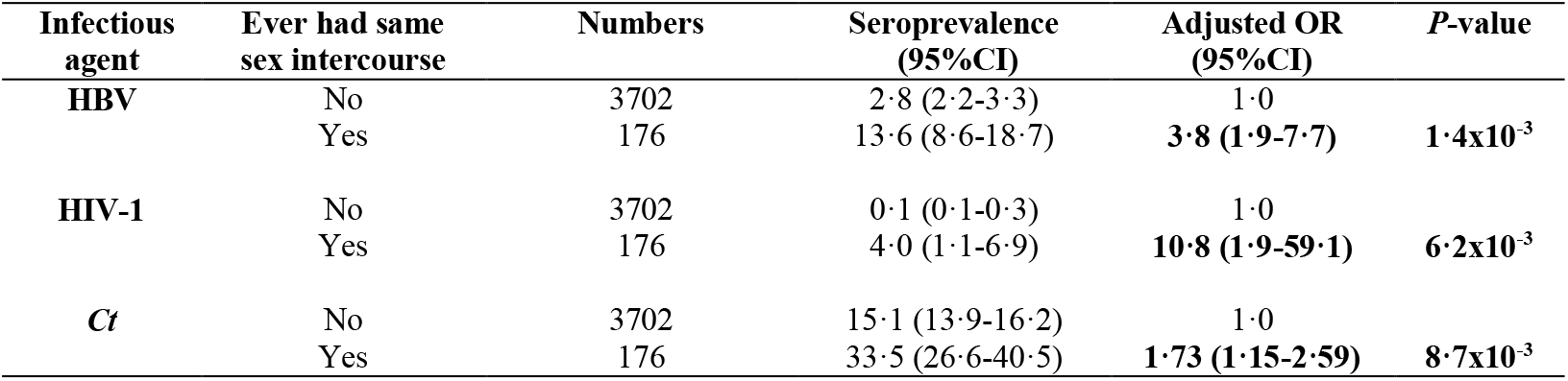
Crude seroprevalence and adjusted odds ratios for HBV, HIV-1 and *Ct* in men reporting same sex intercourse in males. Odds ratios adjusted for age interval, ethnicity, TDI and number of reported lifetime sexual partners.

**Figure 2:**
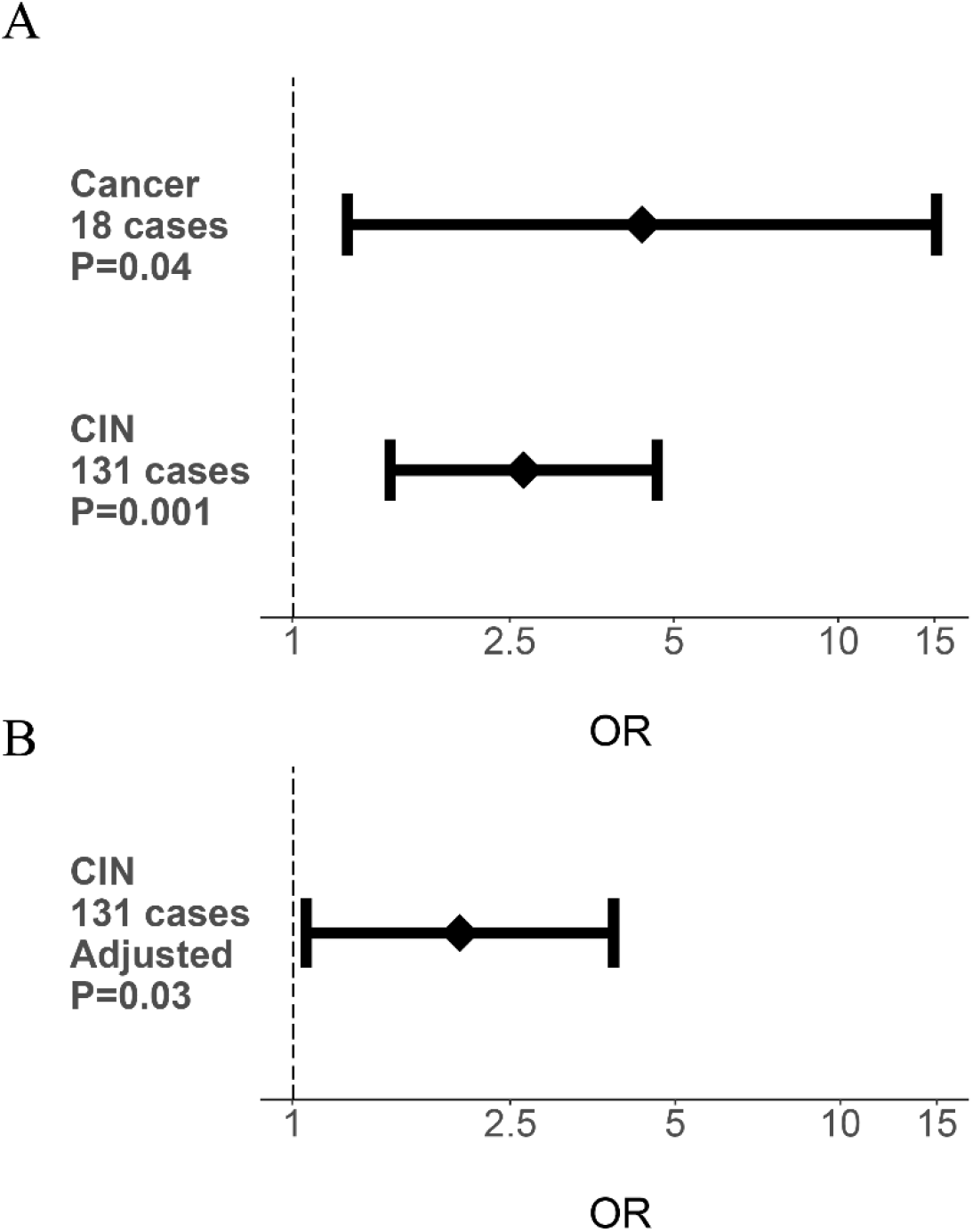
Association between HPV-16 L1 antigen seropositivity and risk of cervical cancer and cervical intraepithelial neoplasia in 9,695 UK Biobank participants. A) Unadjusted odds ratios of cervical cancer and cervical intraepithelial neoplasia (CIN) by HPV-16 L1 seropositivity are shown with 95% confidence intervals (*P*-values calculated with Fisher’s exact test). B) Odds ratio of CIN risk after adjustment for age, sex, ethnicity, LSP and TDI (*P*-value calculated using multivariable regression analysis).

The serum samples were assayed in two equally sized batches, each assayed over a one week period, including blind-spiked duplicates for 107 (1·1%) individuals to quantify within- and between-batch variation (**Supplementary Table 4 and Supplementary Figure 3**). Consistency of serostatus between baseline and repeat assessment samples were tested for 277 individuals to estimate seroconversion and seroreversion rates (**Supplementary Methods and Supplementary Table 5**). Additionally, three reference control sera with known reactivity patterns for the infectious agent panel were tested on each plate to verify uniform plate handling **(Supplementary Methods)**.

**Table 4:**
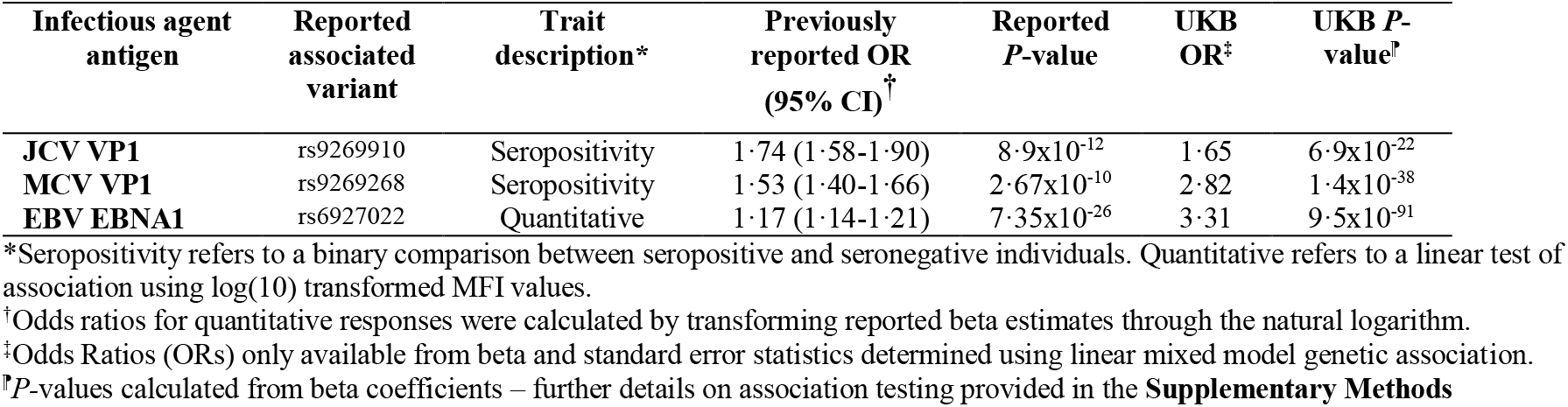
Replication of genetic variants previously associated with exposure to specific infectious agent antigens.

**Figure 3:**
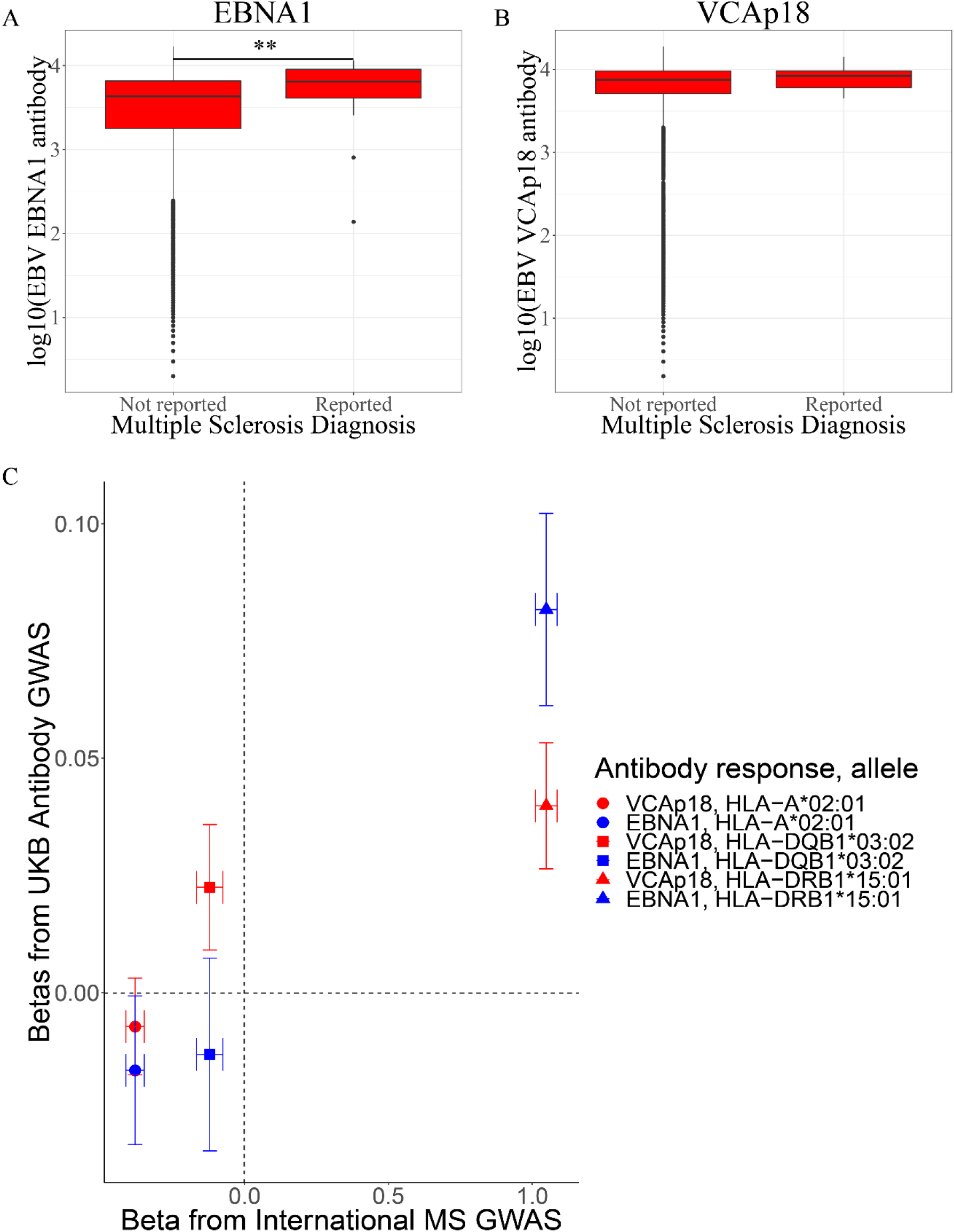
EBV EBNA1 and VCAp18 antibody responses and associations with multiple sclerosis. Log_10_ transformed antibody levels against EBNA1 (A) and VCAp18 (B) antigens were compared between groups in the UKB Multiplex Serology subset with or without self-reported diagnoses of multiple sclerosis (34 cases). **: *P*<0·01. C) The beta coefficients from the GWAS analyses of quantitative antibody responses against EBNA1 (blue) and VCAp18 (red) in the UKB subset (y-axis) were compared to the coefficients from the largest available case-control GWAS of MS (x-axis) for HLA alleles recognised to be associated with MS risk. The points are shaped by imputed HLA allele.

### Statistical analysis

Statistical measures (sensitivity, specificity, Cohen’s *kappa*) for Multiplex Serology assays using reference panel samples^10-12,14,16^ including 95% confidence intervals (CI) were calculated using SAS 9·4.

During Multiplex Serology validation **(Supplementary Figure 1)** we defined cut-off values distinguishing positive versus negative antibody responses against individual antigens based on previously published analyses^10-12,14,16,20-22^. These antigen-specific seropositivity results were subsequently combined using published algorithms to define overall seropositivity for infectious agents (as discussed in more detail on the UK Biobank Data Showcase: https://biobank.ctsu.ox.ac.uk/showcase/refer.cgi?id=1348).

The seroprevalence estimates for each infectious agent were compared to those expected for European populations based on previously published data, determined through a PubMed search using the terms ‘seroprevalence’, ‘prevalence’, ‘IgG’ or ‘antibody’ in addition to the abbreviated or full name of the agent (**Supplementary Methods**).

Analyses were performed that tested associations between seropositivity (for individual antigens) or overall infectious agent seropositivity (i.e. seroprevalence) as well as log_10_ transformed quantitative MFI antibody responses (i.e., seroreactivity) with environmental, demographic and genetic exposures and disease outcomes. The variables used were defined based on existing knowledge of such associations and are described in the **Supplementary Methods**. The variables included sex, age (by decade), ethnicity, Townsend deprivation index (TDI) quintiles, lifetime number of sexual partners (LSP in categories of ‘0’, ‘1’, ‘2-4’, ‘5-10’, ‘Greater than 10’) and self-report of ever same-sex intercourse (sameSI) using a univariate regression model tested against the null (outcome ∼ 1) using the likelihood ratio test (R version 3·5·1). A chi-squared test for trend was used to test for linear upward or downward trends of seroprevalence in the ordered quintiles of TDI. For those variables demonstrating any evidence of association (*P*<0·01), multivariable logistic regression was used to test for association whilst adjusting for other covariates with or without stratification by sex. Similarly, associations of seropositivity with disease prevalence was performed using either univariate logistic regression or, where sufficient power was present, multivariable regression accounting for relevant covariates. Disease outcomes included cervical cancer, multiple sclerosis (MS) and coeliac disease, selected owing to their known (cervical cancer with HPV-16), potential (MS with EBV and CMV exposure) and unlikely (coeliac) association with infection exposure. Disease outcomes were obtained through either self-report, or cancer registry data (details of which are provided in the **Supplementary Methods)**.

### Genetic analysis

Details of the generation of the genotype data have been described elsewhere^23^ and further details are provided in the **Supplementary Methods**. To compare association statistics against genetic association signals for multiple sclerosis, genotypes and case-control phenotypes from the International Multiple Sclerosis dataset and GWAS analysis were used^24^. Human leukocyte antigen (HLA) alleles were imputed using genotype data available from across the extended MHC region only (chromosome 6 base pair positions 25,500,000-34,000,000 in genome build 37) using the SNP2HLA algorithm. Imputation and subsequent association analyses were performed in PLINK 1·9 using logistic regression including sex and age as fixed effect covariates, then statistics from individual countries were meta-analysed using a fixed effects model in METASOFT. Association and bivariate GREML analyses of genetic data were undertaken using the GCTA software (version 1·26·0) for directly genotyped variants. This uses a linear mixed model to control for cryptic relatedness and to allow inclusion of all individuals irrespective of ethnic origin through the calculation of a genetic relatedness matrix (GRM) incorporated as a random effect covariate. BOLT-LMM was an equivalent software used for imputed data. Age and sex were included as fixed effect covariates in all models.

### Data availability

All data are available through the UKB Access Management System (https://bbams.ndph.ox.ac.uk/ams/).

## Results

### UKB Sample Overview

The baseline characteristics of the 9,695 UKB individuals included in the Multiplex Serology study compared with the rest of the cohort with serum available for future analyses are shown in **Table 1**. Of these individuals, 9,611 also had genetic data available for analyses following quality control.

Seroprevalence estimates for each infectious agent, overall and in men and women separately, are provided in **Table 2**. For all infectious agents, seroprevalence estimates were within ranges consistent with those previously reported in European or North American populations.

### Associations between demographic factors and infectious agent seropositivity

We observed differences in seroprevalence estimates between males and females for 11 infectious agents, with 5 found to be higher in women (EBV, HHV-7, HPV-16, HPV-18 and *Ct*) and 6 higher in men (VZV, HBV, HTLV-1, JCV and BKV and *Hp*). For all of the agents where data from multiple antigens were used to calculate total infectious agent seroprevalences, equivalent differences were observed in the same directions for the individual antigens (**Supplementary Table 6**). The crude differences observed by sex for these 11 infectious agents persisted following adjustment for age, TDI, self-reported ethnicity, LSP and sameSI (**Supplementary Table 7**). We also observed significant differences in seroprevalence by age (**Figures 1A and 1B** and **Supplementary Table 8**), being strongest for HSV-1, CMV and *Tg*. Age was inversely associated with HPV-16, JCV and BKV. For example, BKV seroprevalence was 97·0% (95% CI 96·1-97·9%) in men aged 40-49 years, and 93·0% (91·8-94·1%) in 60-69 year olds.

We observed significant differences in seroprevalences of multiple infectious agents by self-reported ethnicity (**Figures 1C and 1D** and **Supplementary Table 9**). For example, there was a higher CMV seroprevalence in Asian (93·6%; 89·6-97·7%) compared to White individuals (56·5%; 55·5-57·5%) and a higher seroprevalence of HBV in Black (23·4%; 16·4-30·4%) compared to White individuals (1·7%; 1·4-1·9%). We also found a higher seroprevalence of several infectious agents known to be transmitted through sexual or close physical contact with increasing LSP (HSV-1, HSV-2, EBV, HPV-16 and HPV-18 and *Ct*; **Figures 1E and 1F** and **Supplementary Table 10**). We did not observe significant associations between LSP and rare sexually transmitted infections such as HIV-1, HBV, HCV or HTLV-1. We did however, observe statistically significant associations in men reporting same sex intercourse for two of these infections (HBV, HIV-1) as well as *Ct* (**Table 3**). We observed an association between higher socio-economic deprivation and many infectious agents including HSV-1, HSV-2, EBV, CMV, KSHV, HBV, *Tg, Hp* and *Ct* (**Supplementary Table 11**). This was particularly striking for *Hp* in women where 25·7% (22·2-29·2%) of women in the least deprived group had evidence of exposure compared to 42·8% (38·6-47·0%) in the most deprived group (*P*_*trend*_=4·3×10^−11^). Similarly, 19·7% (17·4-22·1%) of women in the first quintile had evidence of exposure to *Ct* compared to 34·8% (31·9-37·7%) of women in the fifth quintile (*P*_*trend*_=2·5×10^−16^). The same trends were also observed for males. However, with the exception of *Hp*, none of the associations remained significant after adjusting for age, ethnic group and LSP.

### Genetic associations with infectious agent antibody responses

We replicated three previously reported associations between human genetic variants and antibody responses against infectious agents (**Supplementary Table 12**). All three associations are with variants in the class II region of the MHC locus in the human genome. Two variants (rs9269910 and rs9269268) were found to be associated with seropositivity for JCV (*P*=6·9 x 10^−22^) and MCV (*P*=1·4×10^−38^) respectively, and another variant (rs6927022) was associated with antibody levels against EBV EBNA1 antigen (*P*=9·5×10^−91^; **Table 4**). Furthermore, we discovered a novel, statistically significant signal of association between MHC variants (with the most significant association with the rs7197 variant) and antibody response against the EBV viral capsid antigen (VCAp18; beta 0·04; 95% CI 0·03-0·05; *P*=1·7×10^−22^, **Supplementary Figure 5**).

### Associations between infectious agent exposure and health outcomes

We replicated the well-established association between HPV-16 L1 seropositivity (considered a marker for cumulative exposure) and the risk of cervical intraepithelial neoplasia (CIN; based on 131 cases; OR=2·65; 95% CI 1·51-4·66, *P=*0·001) and cervical cancer (18 cases, OR=4·37; 95% CI 1·26-15·15, *P*=0·04). The association with CIN remained significant after adjustment for age, ethnicity, LSP and TDI (OR=2·02; 95% CI 1·06-3·05; *P*=0·03; **Figure 2**). We found that all self-reported cases of MS disease in the UKB subset (n=34) were seropositive for EBV infection as determined using either our validated algorithm incorporating all four EBV antigens, or using VCAp18 seropositivity alone **(Figure 3A)** which remained significant after adjustment for age, sex, ethnic group and TDI (OR=5·3; 1·55-18·39; *P*=7·8×10^−3^). A similar, non-significant pattern of association was observed for VCAp18 antibody levels **(Figure 3B)**. We also observed an inverse association with CMV seroprevalence and risk of MS that persisted after adjustment for the same covariates (OR=0·39; 0·18-0·81; *P*=0·01).

### Genetic Correlation Analysis

Using imputed HLA allele information we found that several alleles reported to be associated with MS disease risk had similar statistics of association compared to the alleles associated with antibody responses against the EBV antigens EBNA1 and VCAp18 **(Figure 3C)**. To determine the extent of association between EBV antigen response traits and MS risk we calculated the genetic correlation between quantitative VCAp18 and EBNA1 antibody response associations and MS case-control status using the largest available independent case-control analysis of MS. We found statistically significant evidence of a positive genetic correlation between MS risk and VCAp18 antibody response using variants across the MHC region alone (r_G_=0·30, *P*=0·01) and a similar level of correlation using variants across the entire genome (r_G_=0·21, *P*=0·02). We did not observe genetic correlation between EBNA1 antibody levels and MS (r_G_=0·09, *P*=0·41). Using data on coeliac disease (1,468 cases and 10,000 randomly selected controls) within UKB as a negative control we found no evidence of genetic correlation between VCAp18 antibody responses and coeliac disease when tested across the genome or across the MHC (r_G_ = −0·09, *P* = 0·49).

## Discussion

Seven viruses and one bacterial species are established causes of cancer. These and many other species have been hypothesised to contribute to a range of NCDs including cardiovascular and inflammatory conditions. Confirming and understanding the relationships underlying such associations could have significant implications for public health and facilitate the discovery of novel therapeutics. Here we present data from a Multiplex Serology platform applied to a subset of a very large prospective cohort study. Our findings replicate many established relationships, highlighting the validity of our data, and through these results alongside novel associations we demonstrate the potential of the dataset for addressing multiple questions relating to the epidemiology of infectious agents and biology underlying subsequent disease.

Our seroprevalence estimates were consistent with previously published estimates in the UK and Europe. Some of our seroprevalence estimates were on the higher limit of expectations. For example, the *Ct* seroprevalence in women was approximately 25% in our study, and thus significantly higher than in other studies using the microimmunofluorescence (MIF) assay or major outer membrane protein (MOMP) peptide ELISA^16,25^. However, our seroprevalence estimate was consistent with recent data from England based on the same antigen (pGP3) in ELISA assays^26^. This highly immunogenic antigen is believed to be the most species-specific marker of *Ct* that may provide a more accurate estimate of Ct exposure. We have previously demonstrated high agreement of our Multiplex Serology pGP3 assay and the published pGP3 ELISA methods^16^.

Our observations of demographic associations with infectious agent seroprevalence, such as those between LSP and sexually transmitted infections like *Ct* or HPV-16 further enhance the reliability of the seroprevalence estimates derived from our Multiplex Serology panel. Although we did not observe similarly significant associations between LSP and other infections known to be sexually transmitted (HBV, HCV, HIV-1 and HTLV-1), these infections are rare and the existing analysis may be underpowered to detect such associations. Instead, the comprehensive data availability from UKB participants including sensitive aspects of lifestyle meant that we could observe alternative relationships with sexual behaviour reports such as sameSI with HBV, HIV-1 and *Ct* serostatus, thus increasing confidence even in those infections that are rare within UKB.

Many of the demographic associations we observed, such as those with sex, age or deprivation status, have been reported previously but are poorly understood; and such associations may occur as a result of confounding. UKB offers the opportunity to clarify such complex inter-relationships. For example, we were able to demonstrate clear associations with age and CMV^27^, HSV-1^28^ and *Tg*^*29*^ that are likely to be independent and not confounded by other reported risk factors. Such infectious agents have repeatedly been shown to be transmitted through behaviours in adulthood including close physical and sexual contact, and animal exposure or diet as in the case of *Tg*. The associations of CMV, HBV and *Tg* we observed with ethnicity were particularly striking. Although these associations remained highly significant after adjustment for age, sex, LSP and TDI, it is likely that these relationships occur as a combination of environmental and genetic factors in addition to the increased likelihood of infectious agent exposure in other countries where participants of alternative ethnicities may have lived when younger. In contrast, although we observed significant associations between multiple infectious agents and deprivation, these associations were no longer significant after adjustment for age, ethnicity and LSP. Such findings could have significant implications for public health interventions for efforts to improve health equality across strata of society.

The replication of a series of genetic associations with both seropositivity and quantitative antibody responses further increases confidence in the validity of the dataset. Furthermore, the discovery of a new signal of association with antibody responses against EBV VCAp18 highlights the potential of this dataset for novel discovery of genetic variant associations. Associations of pathogen-specific antibodies with the MHC locus are mechanistically highly plausible as variation in *HLA* genes may modulate an individual’s immune response to an infection influencing susceptibility for and the ability to clear infections, and the magnitude of antibody response.

Such genetic associations offer opportunities to improve our understanding of biological mechanisms when interpreted alone. In addition, they can also be used to understand relationships between infections and other NCDs through testing for the presence of shared heritability. This is particularly relevant given the current availability of infectious agent data for only a subset of the existing UKB. Although we had reasonable power within the UKB subset to observe the well-recognised, causative association between HPV-16 seroprevalence and CIN and cervical cancer^30^, testing for associations with small to moderate effect sizes is difficult owing to the limited sample size. Nevertheless, when testing a hypothetical association that has long been speculated to be causal, we did still observe differences in the magnitude and seroprevalence of EBV antibody responses in patients with self-reported MS compared to those without MS in directions consistent with previous reports^5^. By comparing genetic association signals derived from the antibody response in the UKB subset against signals generated from an independently recruited case-control analysis of MS, we observed a significant degree of genetic correlation between EBV VCAp18 antibody responses and MS risk. Our estimation with an r_G_ between 0·2 and 0·3 both across the MHC and the entire genome is in line with previous reports and has been interpreted as evidence in favour of a causal relationship between EBV and MS^31,32^. However, since the majority of this correlation will be driven by the MHC region well known to demonstrate significant levels of pleiotropy (i.e. a single genetic variant may be associated with multiple traits) it is not possible to definitively exclude other confounders that may better explain this relationship. These confounders may indeed include other infectious agents with similar associations across the MHC. Reassuringly we observed no association with coeliac disease that was included as a negative control.

Although we have demonstrated a range of potential utilities for the data generated from the Multiplex Serology panel, it is important to recognise the limitations of our approach. Multiplex Serology is an epidemiological screening tool designed to detect cumulative (i.e. past or present) exposure and thus may be less sensitive or specific than assays used in clinical diagnosis. Moreover, the multiplex nature of the assay does not allow for the optimisation of assay conditions for each and every infectious agent as demonstrated by the variation in validation statistics (e.g. as seen for *Tg*). Nevertheless, even for *Tg*, we were still able to observe robust epidemiological associations consistent with the literature, reinforcing the high quality of the data for epidemiological analyses. Irrespective of all of the above, the availability of additional biomaterials (such as saliva and buffy coats) from baseline and follow-up sampling in the UKB participants offers significant future opportunities. For example, it may be possible to undertake complementary assays (such as nucleotide-based detection systems) to add data about acute infections in individuals identified as seropositive for either prior exposure or carriage using data from the Multiplex Serology panel.

We show that our Multiplex Serology panel offers an attractive validated approach to identify novel and confirm previously reported associations between infectious agents and a range of demographic factors and genetic variants that may influence the risk of chronic disease. In time, with further cohort maturation and investigation by the scientific community, these data are likely to have significant impact on our understanding of the burden of infectious agents and their sequelae, which will in turn inform future public health strategies.

## Research in context

### Evidence before this study

Multiple epidemiological studies have estimated the burden of prior exposure or chronic carriage with a variety of infectious agents. By searching PubMed using criteria as listed in the **Supplementary Methods** we found examples of such cross-sectional studies including the National Health and Nutrition Examination Survey (NHANES), the Detroit Neighbourhood Health Study, the Multi-Ethnic Study of Atherosclerosis and Add Health in the USA, and national surveillance systems implemented in various countries including the UK. Such studies have led to an increased appreciation of disease risk across communities and populations, and in some cases have contributed data in support of infection-disease associations. However, cross-sectional studies cannot determine directionality of such infection-disease associations and are therefore difficult to interpret. Seroepidemiology data from large, broadly characterised prospective cohort studies such as UK Biobank (UKB) and the European Prospective Investigation into Cancer and Nutrition (EPIC) offer opportunities to investigate the temporality of associations between past exposure to infectious agents and subsequent disease risk. Despite the value of such cohort study designs, large-scale serology testing has rarely been undertaken owing to financial and logistical barriers and a tendency to undertake nested case-control analyses to answer specific questions. Furthermore, very few studies have considered infections that are either very rare or very common in populations owing to consequences in terms of statistical power. Some infections, such as HIV-1, have additional ethical barriers to surveillance and detection that confer a risk of selection bias dependent on the populations amenable to study.

### Added value of this study

This study demonstrates the utility of a scalable Multiplex Serology platform to measure antibodies against 45 antigens from 20 infectious agents in a random subset of 9,695 UKB participants. Our seroprevalence estimates fall within expected UK age-sex distributions for all tested infectious agents. The associations observed between seroprevalence and sex, ethnicity and sexual behaviour for multiple infectious agents not only serve to validate the utility of the panel but should also guide public health initiatives to reduce the burden of infections, and thus their immediate symptoms and subsequent disease sequelae. The associations with human genetic variation similarly offer reassurances in terms of cross-cohort reproducibility, and provides an opportunity to discover novel genetic associations, such as that observed between human leukocyte antigen complex variation and EBV VCAp18 antigen responses that offer biological insights into disease pathogenesis research and prevention. If, as planned, data on infectious agents were available for all 500,000 UK Biobank individuals, this resource would represent a unique opportunity to understand how exposure to a range of infectious agents influences subsequent disease risk and how host genetic factors influence these relationships.

### Implications of all the available evidence

The work presented here is the first phase of infectious agent exposure characterisation in UKB and has generated comprehensive data that is now publically available. It represents an exemplar of the types of analyses possible using the wealth of data collected from all UKB participants. The seroprevalence estimates offer opportunities to understand differences by demographics highlighting, for example, the importance of sexual behaviour in disease risk, whilst explaining differences by deprivation status through combinations of age, ethnicity and sexual behaviour. Furthermore, the availability of quantitative seroresponses, as highlighted by the correlation between EBV antigen responses and MS, offers opportunities to estimate the magnitude of disease risk. These investigations will help inform public health initiatives, and generate novel hypotheses for future mechanistic studies.

## Data Availability

All data are available through the UKB Access Management System (https://bbams.ndph.ox.ac.uk/ams/)

https://biobank.ctsu.ox.ac.uk/showcase/refer.cgi?id=1348

## Acknowledgements

We thank the UKB participants and all individuals involved in recruitment, data and sample collection, data curation and release. The authors would like to thank Drs James Gilchrist, Tom Parks and Kathryn Auckland, Mr Andres Kaufmann and Professor Andres Moreno-Estrada for advice and assistance during data analysis. AJM was supported by a Wellcome Trust Fellowship with reference 106289/Z/14/Z and the National Institute for Health Research (NIHR) Oxford Biomedical Research Centre (BRC). The views expressed are those of the authors and not necessarily those of the NHS, the NIHR or the Department of Health. AYC was supported by the Research Councils UK Newton Fund Award with reference MR/N028937/1.

## Author Contributions

AJM, NA, RA, MH, SS, GM, the UKB Infection Advisory Board, RC, AVSH and TW conceptualised and designed the methods for the research project; AJM, NB, TJL, AYC, AC, and RA performed the analyses; AJM, NB, NA, TJL, RA, MH, SS, RC, and TW were responsible for generation of resources and curation of data; AJM, NA, GM, RC, AVSH, and TW were responsible for funding acquisition and supervision, AJM, NB, NA, TJL, RC, AVSH, and TW were responsible for writing the manuscript and all co-authors reviewed the manuscript.

